# Did Risk-based or Age-based Vaccine Prioritization for Covid-19 Save More Lives?

**DOI:** 10.1101/2022.10.18.22281237

**Authors:** Joeri Smits, Amyn A. Malik, Jad A. Elharake, Ahmed Mushfiq Mobarak, Saad B. Omer

## Abstract

**Importance:** All U.S. states provided Covid-19 vaccine access to frontline healthcare workers first, but after that, states varied in whether they gave earlier access to the elderly, versus the vulnerable with comorbidities, or school employees or essential workers, reflecting the underlying scientific and policy uncertainty.

**Objective:** To evaluate if risk-based or age-based prioritization is more effective at reducing reported Covid-19 cases and deaths.

**Design:** A serial cross-sectional study

**Setting:** 50 U.S. states and Washington D.C.

**Participants:** 60+ years of age, high-risk individuals, K-12 school employees, and essential workers

**Main Outcomes and Measures:** Hospitalizations and deaths

**Results:** Seven to nine weeks after 60-year-olds became eligible for a vaccine, there was a statistically significant 40-50% decline in Covid-19 hospitalizations in that state. In contrast, there was no statistically detectable change in hospitalizations in the 7-9 weeks after K-12 employees become eligible for vaccines. Vaccine eligibility of “high-risk adults” and “essential workers” produces effects somewhere in the middle, with reductions in hospitalization of about 25%. There was a large statistically significant decline in death rates (25-38%) 10 to 11 weeks after people aged over 60 became vaccine-eligible. These effects were generally statistically larger than high risk individuals, K-12 school employees, and essential workers.

**Conclusions and Relevance:** Panel data analysis of weekly variation in Covid-19 health outcomes reveals that prioritizing adults 60+ years of age is associated with the largest reduction in hospitalizations and Covid-19 cases, followed by vaccines for adults with high-risk comorbidities. Vaccinations extended to K-12 school employees and essential workers is associated with the smallest reductions in hospitalizations and deaths.

**Key Points:** *Question:* Did Risk-based or Age-based Vaccine Prioritization for Covid-19 Save More Lives?

*Findings:* Panel data analysis of weekly variation in Covid-19 health outcomes reveals that prioritizing adults 60+ years of age is associated with the largest reduction in hospitalizations and Covid-19 cases, followed by vaccines for adults with high-risk comorbidities. Vaccinations extended to K-12 school employees and essential workers is associated with the smallest reductions in hospitalizations and deaths.

*Meaning:* Prioritizing adults 60+ years of age can lead to a higher estimated reduction in hospitalizations and deaths, followed by a strategy of prioritizing adults with high-risk comorbidities. Our findings add to the limited evidence for the roadmap for prioritizing use of Covid-19 vaccines, and help address uncertainties about the relative effectiveness of different vaccine strategies.

## Introduction

As vaccinations against SARS-CoV-2 started to roll out in December 2020 in the United States, initial limited supply necessitated phased allocation. Centers for Disease Control and Prevention (CDC)’s Advisory Committee on Immunization Practices (ACIP) recommended a hierarchical phased approach that considered risk factors, morbidity, and mortality along with social and equity priorities.^1,2^ However, the logistics of implementing such an approach proved cumbersome and many states rolled their vaccine program out with simpler age-based allocation. It was not clear which approach (age-based vs risk-based, or some combination of the two) would result in better outcomes with respect to case notifications, hospitalizations, and deaths. Expert opinions and modeling studies provided some support for each approach.^1,3-5^ A decision-modeling study using electronic health records for simulation predicted that risk-based prioritization could be more effective in preventing deaths and household transmission compared to an age-based approach.^6^

Here we take an empirical approach, utilizing the variation in vaccination policies and prioritization across the 50 U.S. states to evaluate which type of prioritization ultimately proved to be more effective at reducing reported Covid-19 cases and deaths.

## Methods

The unit of observation is a U.S. state in a week in our analysis. With 50 U.S. states plus Washington DC, we have 51 serial cross-sectional observations. We have 39 weeks of observations for each of those 51 cross-sectional units.

The three dependent variables are the numbers of cases, hospitalizations, and deaths in a US state in a week. Covid-19 case and death count data are from the Oxford Covid-19 Government Response Tracker (Hale et al., 2020). Covid-19 hospitalization data comes from U.S. Department of Health & Human Services (2022). We collapsed the data from daily to weekly as there was lower reporting on weekends, and there was ex-post corrections to the data leading to instances of negative daily case and death counts. The first US case was detected in week 4 of 2020 and by week 12 of 2020, all 51 U.S. states had registered at least one Covid-19 case. The first reported Covid-19 death was in Washington state in week 9 of 2020. By week 13 of 2020, only four states (Hawaii, South Dakota, Virgin Islands and Wyoming) had not had a single Covid-19 death yet.

The key independent variables of interest are indicator variables for whether a demographic group is eligible to get vaccinated in that week. The indicator for a group is set to 1 in a week if on at least 4 days in that week the group is eligible for vaccination (and it is set to 0 otherwise). The demographic groups are

- Adults aged 60 and over
- Frontline essential workers
- K-12 employees
- Adults with high-risk medical conditions

The eligibility start dates for these groups, with the exception of adults with high-risk medical conditions, is drawn from Covid-19 US State Policies (CUSP) dataset (Raifman et al. 2020). We compiled the dates at which adults with high-risk medical conditions became eligible for vaccination from the websites of health departments and news reports in each state. Frontline essential workers are essential workers that are most likely at highest risk for work-related exposure to SARS-CoV-2 because their work-related duties must be performed on-site and involve being in close proximity (<6 feet) to the public or to coworkers. This group is proxied by grocery store workers, and includes firefighters, law enforcement and public safety personnel, correctional staff, U.S. Postal Service workers, food and agricultural workers, and manufacturing workers. For the eligibility start date of K-12 employees in each state, we conducted a search of states’ public health departments and local newspapers.

### Statistical Analysis

To provide a sense of the variation in vaccine prioritization strategies pursued by different U.S. states, Fig. 1 displays the Covid-19 eligibility start dates for the ACIP-defined groups for the 4 most populous U.S. states.^7^ Fig. S1-S5 in the Supplement show the timelines for all other U.S. states and the District of Columbia. Frontline healthcare workers, emergency medical service providers, and long-term care residents became eligible first in all states, on December 14^th^ of 2020. Thereafter, the prioritization varied by state. Certain states prioritized the vaccination of ACIP-defined groups for the purposes of the recovery of their state economy (e.g., frontline essential workers, and K-12 employees). Other states’ authorities prioritized vulnerable groups (e.g., the elderly, individuals with high-risk comorbidities). For example, K-12 school employees in New York were eligible to receive the vaccine over a month before New Yorkers with high-risk medical conditions, while the reverse was true in Texas (Fig. 1). More generally, the lag in eligibility of specific age groups (60+, 50+) relative to frontline healthcare workers varied across states, and our empirical strategy will exploit such variation to evaluate the effectiveness of alternative strategies. The heatmap in Fig. 2 displays the prioritization order in each state. Table S1 in the Supplement tabulates the relative subpopulation by state.

**Fig. 1:**
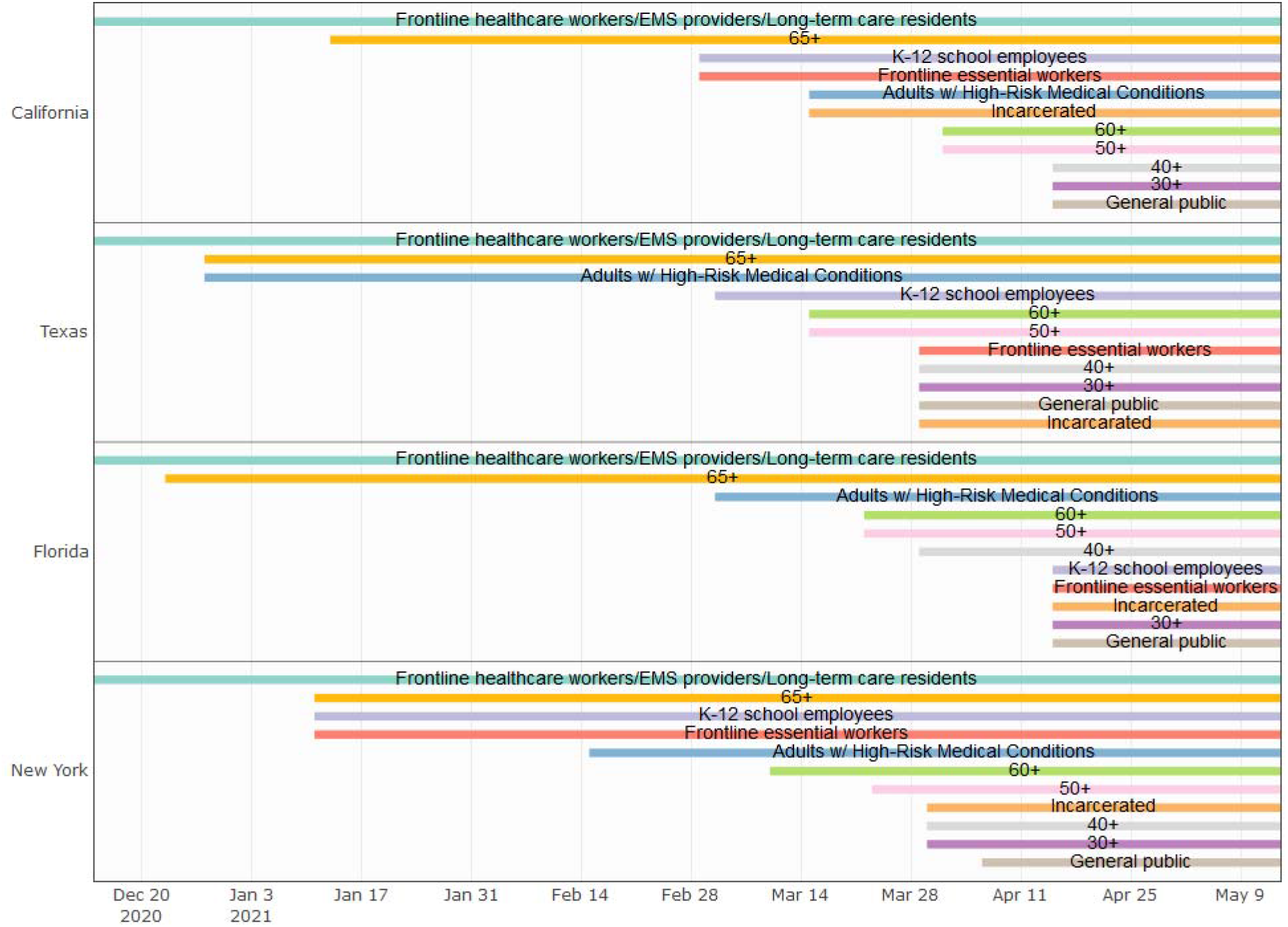
The timing of eligibility for Covid-19 vaccination in the four most populous states of the U.S. Each of these four states started their vaccination campaign on 14 December 2020 by vaccinating frontline healthcare workers, emergency medical service (EMS) providers, and long-term care residents. The ‘65+’ label corresponds to adults ages 65+ (with analogous labels for the other age groups). The eligibility start date was the same for the 65+, 70+, 75+ and 80+ groups in the four states shown. ‘Frontline essential workers’ are essential workers that are most likely at highest risk for work-related exposure to SARS-CoV-2, because their work-related duties must be performed on-site and involve being in close proximity ($<$6 feet) to the public or to coworkers. This group is proxied by grocery store workers, and includes firefighters, law enforcement and public safety personnel, correctional staff, U.S. Postal Service workers, food and agricultural workers, and manufacturing workers.

**Fig. 2:**
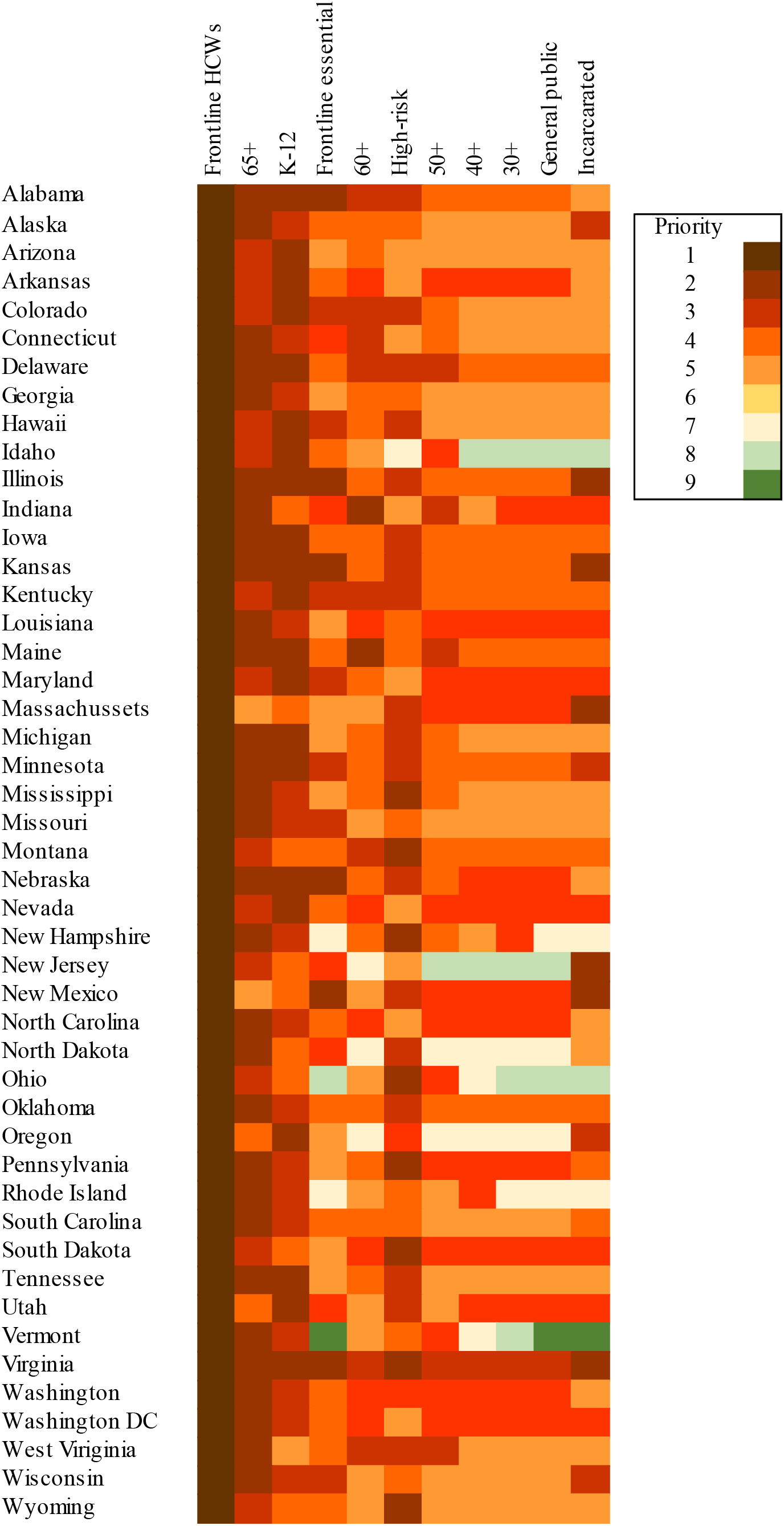
Heatmap of the order in which groups became eligible for Covid-19 vaccination, across states. Our analysis focused on the effects of the variation in vaccine eligibility dates for the four ACIP-defined groups for whom there was meaningful variation across states: “Population aged 60+”, “Adults with high-risk medical conditions”, “Frontline essential workers”, and “K-12 employees.”^7^ We regress Covid-19-related hospitalizations in week *w* in state *s* on whether each of these four groups of interest were eligible to receive a vaccine in week (*w-7*) in that state. We choose a 7-week lag structure because we expect hospitalizations to respond to changes in vaccination with at least a 7-week lag. We explore the sensitivity of our results to changing this lag structure to 8, 9, 10, 11, or 12 weeks. Covid-19 hospitalizations is measured as the number of patients who were admitted to an adult inpatient bed on the previous day who had Covid-19 at the time of admission in the state. These data were drawn from the U.S. Department of Health & Human Services.^8^

States and their governors may have possessed differing beliefs about the effects of various vaccination prioritization schemes on Covid-19 burden of disease. The relative population size of the ACIP-defined groups (e.g., elderly share of the population) or the strength of interest groups (e.g., unions) may vary across states. To the extent that these factors may also affect health outcomes, including Covid-19 outcomes and independent of Covid-19 vaccination status, this would pose a threat to the internal validity of our results. However, we analyzed weekly variation in Covid-19 outcomes, controlling for a set of indicator variables for every state that appears in our analysis. The inclusion of state fixed effects in our regressions accommodates any such time-invariant (over the time span of our data) unobserved heterogeneity across states.

We also estimate the effect of variation in vaccine eligibility on Covid-19-related cases and deaths as the outcome variables. We study deaths with a 10-week lag as Covid-19 deaths have typically lagged hospitalizations with a further three weeks. For Covid-19 cases, we use a shorter 6-week lag structure.

We further include the following time-varying controls for each state-week: the number of Covid-19 diagnostic tests conducted, the total number of Covid-19 vaccines administered, a variable counting the number of weeks since the week wherein individuals aged 80+ became eligible for vaccination, a non-pharmaceutical stringency index (NPI) capturing restrictions on movement and mask mandates, and a polynomial of third degree of a count variable that takes on 1 for week 13 in 2020 (by this week, only four states (Hawaii, South Dakota, Virgin Islands and Wyoming) had not had a single Covid-19 death yet), to control for common time trends.

## Results

We report linear regressions with log (hospitalizations) as the outcome variable in the odd-numbered columns of Table S2 and Fig. 3, and report Poisson regressions on hospitalization in the even numbered columns, to account for the count-data nature of the dependent variable, and explore sensitivity of results to this modeling choice.

**Fig. 3:**
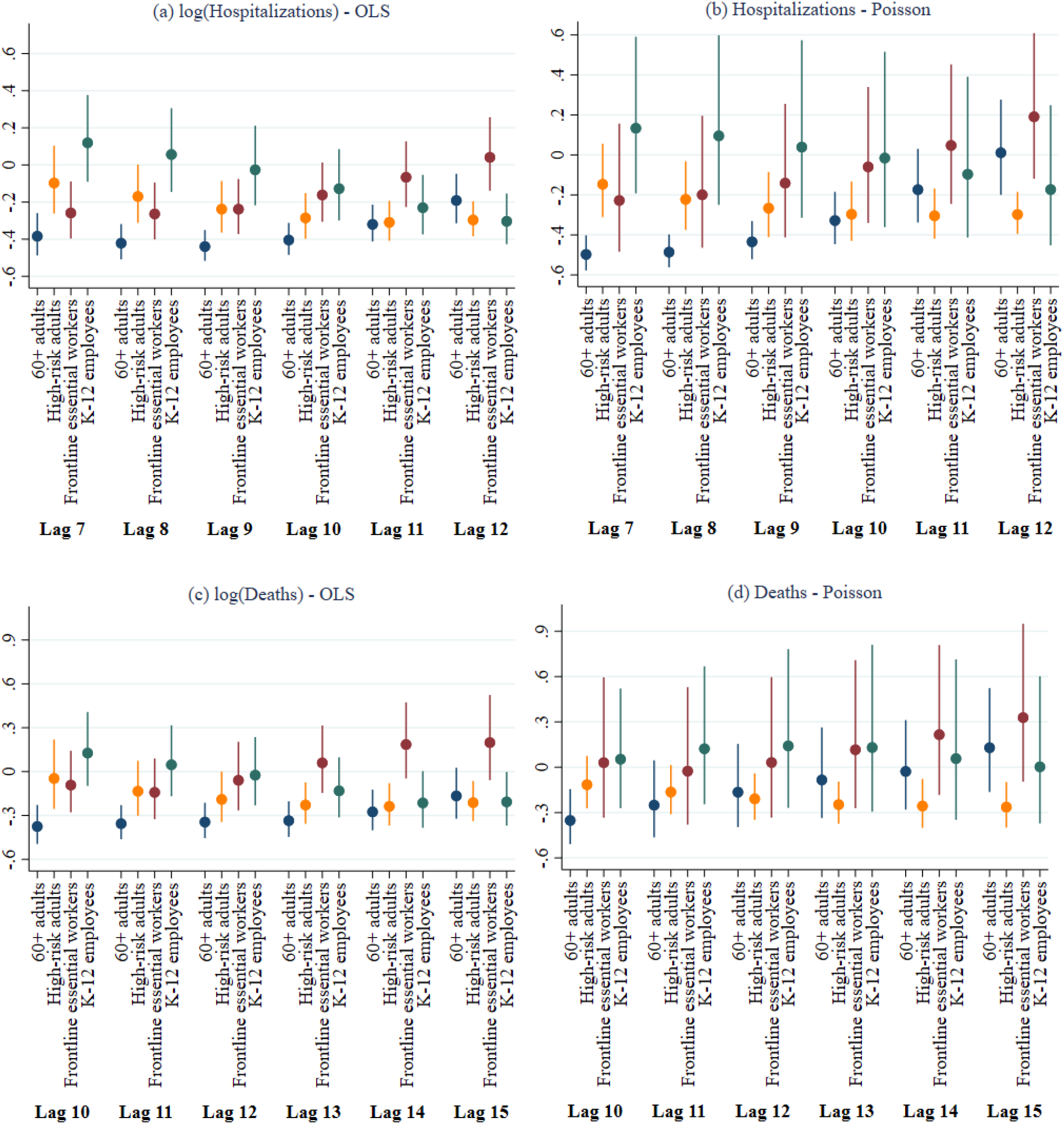
In regressions of Covid-19 hospitalizations and deaths, the coefficients are plotted of the indicators for whether an ACIP group was eligible in a state, with a lag of 7-12 weeks for hospitalizations (panels (a) and (b)) and 10-15 weeks for deaths (panels (c) and (d)). Panels (a) and (c) display estimates of the coefficients and 95% confidence intervals of log-linear models estimated by OLS (after the transformation (exp(β) - 1)), and panels (b) and (d) display coefficient estimates and 95% confidence intervals of Poisson regressions. For all four panels, 100 times the displayed (transformed) coefficients can be interpreted as the percentage change relative to the benchmark where neither of the four groups is eligible for vaccination in that week/lag.

The hospitalization regressions (summarized in Fig. 3, panel (a) and (b)) showed a very clear pattern, where age-based prioritization of people 60 and older is associated with the largest reductions in Covid-19-related hospitalizations 7-9 weeks later. Seven to nine weeks after 60-year-olds became eligible for a vaccine, there was a statistically significant 40-50% decline in Covid-19 hospitalizations in that state. In contrast, there was no statistically detectable change in hospitalizations in the 7-9 weeks after K-12 employees become eligible for vaccines. This was striking because most U.S. states, including some of the largest ones (New York, California, Texas) extended vaccine eligibility to K-12 employees before people aged 60+. In New York, people aged 60+ received vaccines almost two months after K-12 employees.

Vaccine eligibility of “high-risk adults” and “essential workers” produces effects somewhere in the middle, with reductions in hospitalization of about 25%. The larger effect of prioritizing 60-year-olds is statistically distinguishable from the close-to-null effect of prioritizing K-12 employees (p-value<0.001, see Table S2), and also from the smaller effect of making high-risk adults vaccine-eligible (p-value<0.02).

Looking across the different lag structures, we saw that the effect of making adults with high-risk comorbidities vaccine-eligible on hospitalization rates became larger and more statistically precise with longer lags. Hospitalizations were reduced by about 30% ten to twelve weeks after high-risk adults become eligible for vaccination. In contrast, the negative effect of vaccinating frontline essential workers on hospitalizations tapers off after about 10 weeks.

Panel (c) and (d) of Fig. 3 and Table S3 in the Supplement display effects of vaccine-eligibility dates of these four groups on Covid-19-related *deaths* 10-15 weeks later. Again, there was a large statistically significant decline in death rates (of 25-38%) ten to eleven weeks after people aged over 60 became vaccine-eligible. These effects were generally statistically larger than the other three categories. However, when we moved to longer lags (12-15 weeks), the effects were smaller and less precise, especially under the Poisson specification. The linearity assumption appeared more consequential for deaths, perhaps because it is a rarer event than hospitalizations or cases. In contrast, when we moved to longer lags, there was a precise 21-26% reduction in deaths 12-15 weeks after high-risk adults are made vaccine-eligible.

Fig. S6 and Table S4 in the Supplement displays effects on Covid-19 cases 6-11 weeks after vaccine-eligibility status of each of the four groups. Again, age-based prioritization (of 60+) is associated with large, statistically precise, and robust reduction in the Covid-19 caseloads. There is a 50-70% reduction in Covid-19 cases 6-9 weeks after vaccines are extended to people aged over 60. This reduction is statistically larger than the effect of extending eligibility to K-12 employees (p-value<0.006), high-risk adults (p-value<0.008), and essential workers (p-value<0.1 in most specifications). Based on effect size, statistical precision, and robustness to different estimation methods and lag structures, age-based prioritization again appears to be a clear winner, for minimizing case rates.

A potentially important confounder that may plausibly not have varied substantially over the time span of our analysis, is the relative size of each subpopulation (60+ years of age, high-risk individuals, K-12 school employees, and essential workers) in each state. Table S5 in the Supplement regresses indicators for a population group having (possibly shared) top priority among the four groups in a state, on four variables capturing the population sizes of these four groups in the state. The results suggest that the sizes of these populations had no predictive power with respect to whether a state prioritized that group for Covid-19 vaccination.

## Discussion

Our empirical results using variation in Covid-19 vaccine prioritization policies across the U.S. states showed that an age-based prioritization approach targeting those aged 60+ years is associated with larger reductions in hospitalizations (by about 50%), deaths (by 20-40%), and cases (by 50-70%). Further, this appeared statistically more effective than prioritizing frontline essential workers and K-12 school employees. The data suggests that the second-best strategy, after providing vaccines to the elderly, is to prioritize high-risk individuals with co-morbidities. Vaccine eligibility for high-risk individuals was associated with a 20-30% reduction in hospitalizations and a 20-25% reduction in deaths. For the “death” outcome, prioritizing high-risk individuals was as effective as prioritizing those aged 60+.

Bubar et al (2021) used a mathematical model to predict the likely effects of age-stratified prioritization strategies, and predicted that mortality would be minimized when people over age 60 are prioritized for vaccines. Our empirical results are consistent with their predictions.^5^ Tran et al (2021) also used a mathematical model matched to prevailing epidemic trends in two U.S. states in early 2021 and proposed prioritizing the elderly.^4^ However, based on computer simulation modeling, Kipnis et al (2021) proposed that a risk-based approach would be better than an age-based approach to minimize hospitalizations ^6^. The empirical data we report, looking retrospectively at actual hospitalization rates and case counts, run counter to this prediction.

Wrigley-Field et al (2021) noted that Covid-19 mortality is substantially higher among Black, Indigenous, and People of Color (BIPOC) populations in the U.S., and vaccine prioritization strategies that target BIPOC can therefore out-perform simple age-based prioritization.^3^ We were unable to directly test this hypothesis because the prioritization strategies used by U.S. states were generally not sensitive to people’s ethnicity, so there was no such variation in our data. But this is also not a confounder to our analysis, because state fixed effects account for cross-state variation in racial composition.

National Academies of Science, Engineering and Medicine (NASEM) used ethical and equitable reasoning in formulating their strategy for vaccine distribution with a goal to “reduce severe morbidity and mortality and negative societal impact due to the transmission of SARS-CoV-2.”^9^ The NASEM framework prioritized healthcare workers, frontline workers, people with high-risk conditions and older adults in congregate settings in Phase 1.^9^ K-12 teachers and other older adults were placed in Phase 2.^9^ Our study showed that prioritizing frontline essential workers and K-12 school employees reduces hospitalizations and deaths to a lesser extent compared to an age-based prioritization scheme or a prioritization of adults with high-risk conditions. An age-based prioritization, similar to that enacted by the United Kingdom,^10^ seems most effective at reducing hospitalizations and deaths, and this approach is also cleaner and easier to implement. Many states ultimately abandoned their plans for a more complicated phased approach in favor of a simpler age-based approach.

Limitations of our study include not being able to estimate effectiveness at a more granular level than state as prioritization policies were mostly implemented at the state level. We were unable to compare strategies targeting healthcare workers against other strategies, as healthcare workers were universally prioritized across the states, leaving no such variation for constructing that test.

A key concern for inference was the possibility that vaccine prioritization of a group reflected the broader clout that the same group enjoys in that state. For example, some groups (e.g., K-12 school employees) may be better organized and have more influence on policy decisions in some states. In that case, the well-organized group that gets vaccine priority may independently enjoy better health through other favored policies. To mitigate these concerns, we include state fixed effects, which filter out any unobserved time-invariant heterogeneity between states. To the extent that potential state-level confounders (e.g., voter blocks’ influence on policy) do not vary over the time span of our data, the inclusion of fixed effects would mitigate such sources of bias in our estimates. Given the speed of Covid-19 vaccine distribution, the relevant variation in our data only spans a few weeks, so it is reasonable to assume that other policies were held fixed during this period. Even after the inclusion of state fixed effects and other time-varying controls like the introduction of non-pharmaceutical interventions in various states, if there are time-varying unobserved factors associated with some states’ decisions to prioritize the elderly rather than K-12 employees (or any other category), that could confound our conclusions. For example, if the elderly or school employees became better organized, or evolved to have stronger or weaker lobbying power in specific states, that could lead them to become vaccine-eligible earlier, and also have independent effects on their health outcome. Our use of weekly variation, and our ability to analyze Covid-19-specific hospitalizations and deaths (as opposed to people’s general health outcomes) in the specific weeks after vaccine-eligibility changes do mitigate these concerns somewhat. Our robustness check of regressing prioritization strategy against share of population in each state also confirmed that this may not influence our results.

A strength of our study is that we took an empirical approach that exploits variation in state prioritization schemes in real-world settings to compare alternative prioritization strategies. This is an important and necessary complement to previously published modeling and simulation approaches, and our analysis produces rigorous empirical tests of the predictions generated by that literature. Further, we explored robustness to various model specifications and lag structures to build confidence in our conclusions.

In this empirical study of COVD-19 vaccine prioritization schemes across the US, prioritizing adults 60+ years of age was associated with highest estimated reduction in hospitalizations and deaths, followed by a strategy of prioritizing adults with high-risk comorbidities. Prioritizing frontline workers, K-12 school employees led to the lowest reduction in hospitalizations and deaths but might have other societal benefits in terms of return to in-person education. Our findings add to the limited evidence for the roadmap for prioritizing use of Covid-19 vaccines, recommended by the World Health Organization’s Strategic Advisory Group of Experts (SAGE),^11^ and help address uncertainties about the relative effectiveness of different vaccine strategies. Although the current vaccine supply is plentiful in the U.S. with no prioritization necessary, findings from this study can inform policies in other countries still rolling out vaccine doses, and can inform future vaccine allocations everywhere.

## Data Availability

All data produced in the present study are available upon reasonable request to the authors.

## Author Contributions

Conceptualization: JS, AAM, AMM, SBO; Methodology and Analyses: JS, AAM, JAE; Project administration: JS, AAM, JAE; Supervision: AMM, SBO; Writing – original draft: JS, AAM, AMM; Writing – review & editing: JS, AAM, JAE, AMM, SBO

## Data and Materials Availability

The researchers on this project will make the data and associated documentation available upon request.

## Supplement

### Material and Methods

We ran panel regressions of dependent variable *y*_*it*_ (case, hospitalizations, or death count) for U.S. state/territory *i* in week *t*

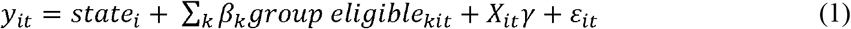

on *group eligible*_*kit*_, which takes on 1 if demographic group

*k* ∈ {*adults aged* 60+, *frontline essential workers, K* − *12 employees, adults w/high* − *risk medical conditions*} is eligible for vaccination in state *i* for at least 4 days in week *t* (and it takes on 0 otherwise), *X*_*it*_ - a vector of time-varying control variables; *state*_*i*_ are state fixed effects and is *ε*_*it*_ is an idiosyncratic error. Standard errors are clustered at the U.S. state level. The independent variables are lagged by 6 to 11 weeks for cases, 7 to 12 weeks for hospitalizations, and 10 to 15 weeks for deaths in the analysis to account for vaccine administration, reporting delays and biological processes.

The control variables include:

- The number of Covid-19 tests administered in state *i* in week *t*, sourced from the JHU Coronavirus Resource Center.
- The number of Covid-19 vaccine doses administered in state *i* in week *t*, sourced from the CDC through Ourworldindata (Mathieu et al. 2021)
- A non-pharmaceutical interventions (NPI) stringency index, sourced from the Oxford Covid-19 Government Response Tracker (Hale et al., 2020).
- A variable counting the number of weeks since 80+ individuals became eligible for vaccination
- A polynomial of third degree of a count variable that takes on 0 for week 13 in 2020, 1 for week 14 in 2020, and so on, to control for common time trends.

Vaccination data by U.S. state are available from Week 2 of 2021. The estimation sample is thus restricted from week 2 of 2021 until week 40 of 2021. Week 2 of 2021 starts on Wednesday January 8^th^, 2021 and ends on January 14^th^, 2021. Week 40 of 2021 starts on October 1^st^, 2021 and ends on October 7^th^, 2021. This leaves 51 states × 39 weeks = 1,989 state-week observations.

The dependent variable is a count, but it contains large values and has only one zero for cases and no zeros for deaths in the estimation sample, so we run both Poisson regressions and ordinary least squares (OLS) regressions with log-transformation of specification (1). Though these variables display overdispersion, the advantage of Poisson is that from pseudo-maximum likelihood (ML) theory, estimates are consistent without the need for a correctly specified likelihood function; only the conditional mean must be exponential (Gourieroux et al., 1984). Thus, the Poisson pseudo-ML estimates are consistent even in the presence of overdispersion. We report in the Tables and Figures in this article the transformed coefficients (exp(*β*) – 1), such that the estimates of the linear models and the Poisson models have the same interpretation: 100 × *β*_*k*_ is the percentage changed in the outcome associated with the vaccination eligibility status of ACIP group *k* changing from non-eligible to eligible. The base (left-out) category in each of the regression is frontline healthcare workers/emergency medical services/long-term care residents. Thus, (100×) the coefficient on an ACIP group indicator captures the percentage change in the outcome corresponding to a change in the vaccination eligibility for that ACIP group, relative to only the frontline healthcare workers/emergency medical services/long-term care residents being eligible for vaccination.

- The data sources for the relative sizes of the subpopulations by state stem from: Share of 60+ adults: the US Census Bureau;
- Share of K-12 employees: the U.S. Bureau of Labor statistics (www.bls.gov/oes/);
- Share of individuals with high-risk co-morbidities: Adams et al. (2020).
- Share of essential workers: U.S. Bureau of Labor Statistics (BLS) and the U.S. Cybersecurity and Infrastructure Agency (CISA).

**Table S1:**
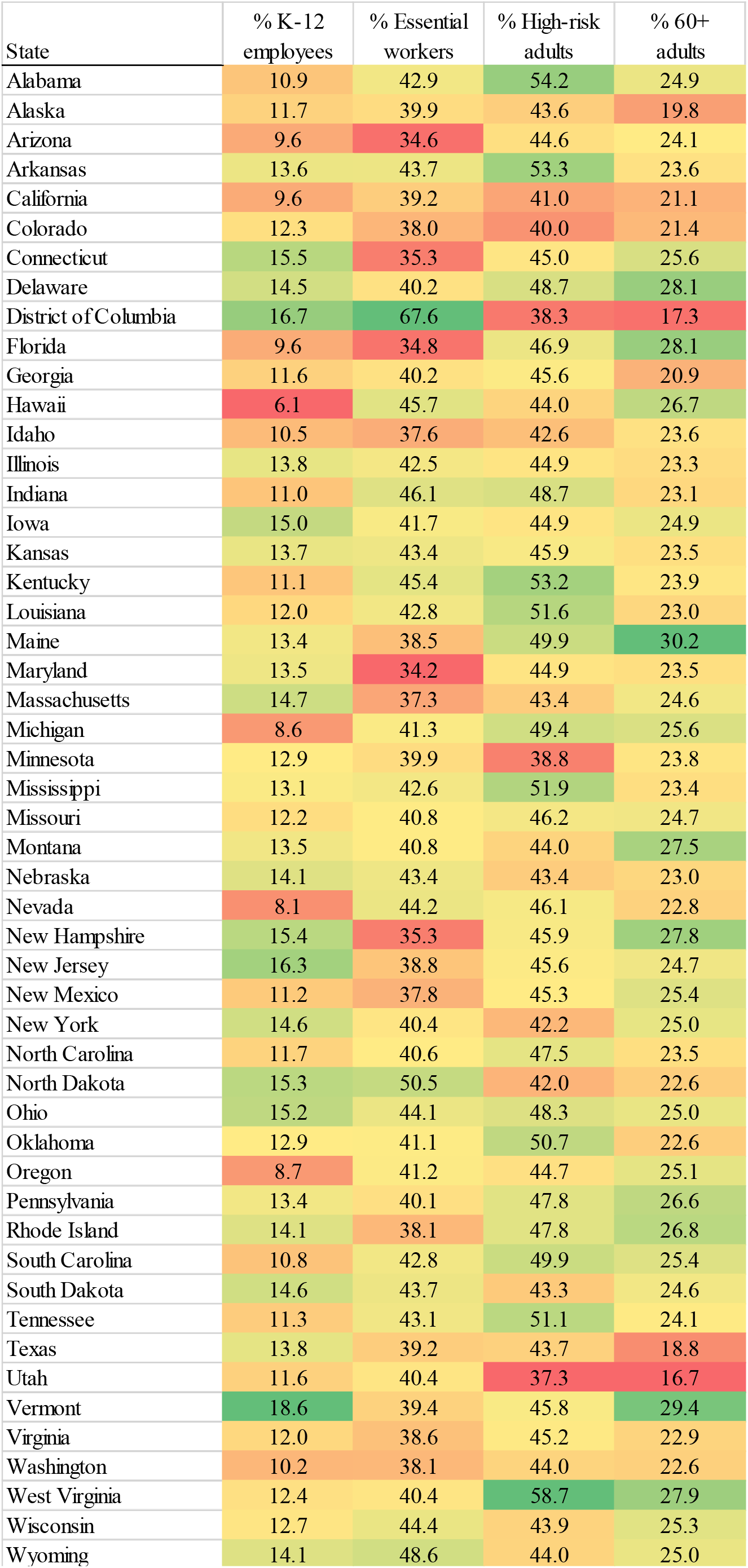
Estimated shares of the four subpopulations by U.S. state.

**Table S2:** Outcome: log(hospitalizations) in odd-numbered columns; hospitalizations (100s) in even-numbered columns.

|  | OLS<br>(1) | Poisson<br>(2) | OLS<br>(3) | Poisson<br>(4) | OLS<br>(5) | Poisson<br>(6) | OLS<br>(7) | Poisson<br>(8) | OLS<br>(9) | Poisson<br>(10) | OLS<br>(11) | Poisson<br>(12) |
| --- | --- | --- | --- | --- | --- | --- | --- | --- | --- | --- | --- | --- |
| 60+ adults (lag 7) | -0.38***<br>(0.09) | -0.50***<br>(0.09) |  |  |  |  |  |  |  |  |  |  |
| High-risk adults (lag 7) | -0.10<br>(0.10) | -0.15<br>(0.11) |  |  |  |  |  |  |  |  |  |  |
| Essential workers (lag 7) | -0.26***<br>(0.10) | -0.23<br>(0.21) |  |  |  |  |  |  |  |  |  |  |
| K-12 employees (lag 7) | 0.12<br>(0.10) | 0.13<br>(0.17) |  |  |  |  |  |  |  |  |  |  |
| 60+ adults (lag 8) |  |  | -0.42***<br>(0.08) | -0.49***<br>(0.08) |  |  |  |  |  |  |  |  |
| High-risk adults (lag 8) |  |  | -0.17*<br>(0.09) | -0.22**<br>(0.11) |  |  |  |  |  |  |  |  |
| Essential workers (lag 8) |  |  | -0.26***<br>(0.10) | -0.20<br>(0.20) |  |  |  |  |  |  |  |  |
| K-12 employees (lag 8) |  |  | 0.06<br>(0.11) | 0.10<br>(0.19) |  |  |  |  |  |  |  |  |
| 60+ adults (lag 9) |  |  |  |  | -0.44***<br>(0.07) | -0.43***<br>(0.09) |  |  |  |  |  |  |
| High-risk adults (lag 9) |  |  |  |  | -0.24***<br>(0.09) | -0.27***<br>(0.11) |  |  |  |  |  |  |
| Essential workers (lag 9) |  |  |  |  | -0.24***<br>(0.10) | -0.14<br>(0.19) |  |  |  |  |  |  |
| K-12 employees (lag 9) |  |  |  |  | -0.03<br>(0.11) | 0.04<br>(0.21) |  |  |  |  |  |  |
| 60+ adults (lag 10) |  |  |  |  |  |  | -0.40***<br>(0.07) | -0.33***<br>(0.10) |  |  |  |  |
| High-risk adults (lag 10) |  |  |  |  |  |  | -0.29***<br>(0.09) | -0.30***<br>(0.11) |  |  |  |  |
| Essential workers (lag 10) |  |  |  |  |  |  | -0.16*<br>(0.09) | -0.06<br>(0.18) |  |  |  |  |
| K-12 employees (lag 10) |  |  |  |  |  |  | -0.13<br>(0.11) | -0.02<br>(0.22) |  |  |  |  |
| 60+ adults (lag 11) |  |  |  |  |  |  |  |  | -0.32***<br>(0.07) | -0.17*<br>(0.11) |  |  |
| High-risk adults (lag 11) |  |  |  |  |  |  |  |  | -0.31***<br>(0.08) | -0.30***<br>(0.09) |  |  |
| Essential workers (lag 11) |  |  |  |  |  |  |  |  | -0.07<br>(0.09) | 0.05<br>(0.17) |  |  |
| K-12 employees (lag 11) |  |  |  |  |  |  |  |  | -0.23**<br>(0.10) | -0.10<br>(0.22) |  |  |
| 60+ adults (lag 12) |  |  |  |  |  |  |  |  |  |  | -0.19**<br>(0.08) | 0.01<br>(0.12) |
| High-risk adults (lag 12) |  |  |  |  |  |  |  |  |  |  | -0.30***<br>(0.07) | -0.30***<br>(0.08) |
| Essential workers (lag 12) |  |  |  |  |  |  |  |  |  |  | 0.04<br>(0.09) | 0.19<br>(0.15) |
| K-12 employees (lag 12) |  |  |  |  |  |  |  |  |  |  | -0.30***<br>(0.10) | -0.17<br>(0.21) |
| F-tests equality of coefficients: |  |  |  |  |  |  |  |  |  |  |  |  |
| 60+ = essential | 0.218 | 0.078 | 0.086 | 0.061 | 0.014 | 0.064 | 0.003 | 0.125 | 0.007 | 0.268 | 0.047 | 0.409 |
| 60+ = K-12 | 0.000 | 0.000 | 0.000 | 0.000 | 0.000 | 0.006 | 0.004 | 0.099 | 0.307 | 0.700 | 0.207 | 0.384 |
| 60+ = high-risk | 0.017 | 0.001 | 0.011 | 0.007 | 0.018 | 0.109 | 0.134 | 0.789 | 0.891 | 0.307 | 0.230 | 0.022 |
| essential = K-12 | 0.015 | 0.247 | 0.031 | 0.364 | 0.131 | 0.593 | 0.807 | 0.899 | 0.228 | 0.674 | 0.014 | 0.272 |
| essential = high-risk | 0.211 | 0.629 | 0.442 | 0.890 | 0.998 | 0.424 | 0.266 | 0.118 | 0.033 | 0.019 | 0.004 | 0.001 |
| high-risk = K-12 | 0.185 | 0.249 | 0.124 | 0.203 | 0.115 | 0.228 | 0.186 | 0.252 | 0.427 | 0.350 | 0.933 | 0.524 |
| Observations | 1,989 | 1,989 | 1,989 | 1,989 | 1,989 | 1,989 | 1,989 | 1,989 | 1,989 | 1,989 | 1,989 | 1,989 |
| (Pseudo-)R<sup>2</sup> | 0.566 | 0.499 | 0.577 | 0.499 | 0.584 | 0.498 | 0.579 | 0.496 | 0.569 | 0.495 | 0.559 | 0.494 |

Note: Robust standard errors clustered at the state level are displayed in parentheses below each respective coefficient, \*\*\* p<0.01, \*\* p<0.05, \* p<0.1 The interpretation of $100 \times \beta$ is the percentage change in hospitalizations in the state-week associated with a change of the regressor from 0 to 1.

**Table S3:**
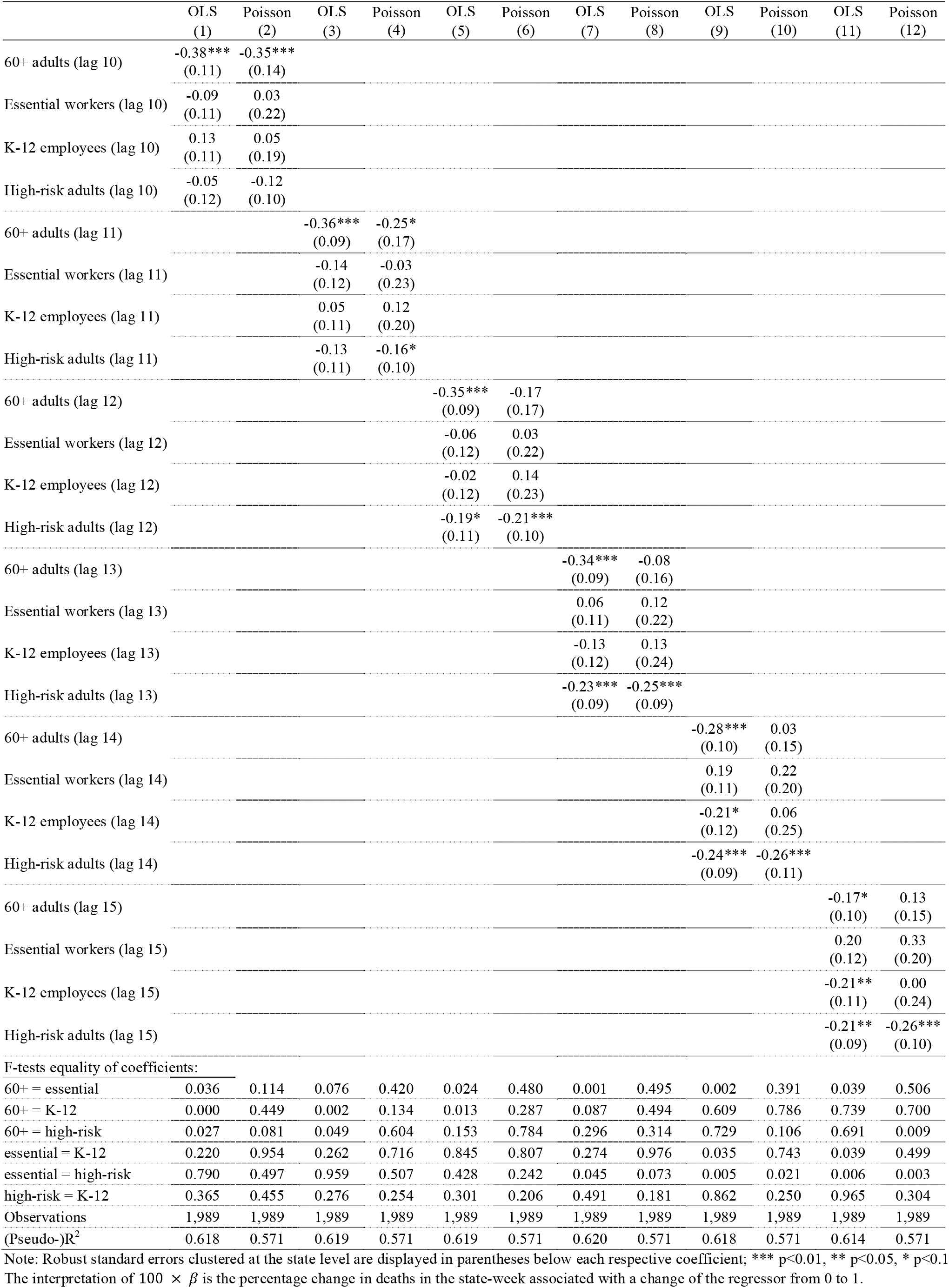
Outcome: log(deaths) in odd-numbered columns; deaths (in 100s) in even-numbered columns.

**Table S4:** Outcome: log(cases) in odd-numbered columns; cases (in 1000s) in even-numbered columns.

|  | OLS<br>(1) | Poisson<br>(2) | OLS<br>(3) | Poisson<br>(4) | OLS<br>(5) | Poisson<br>(6) | OLS<br>(7) | Poisson<br>(8) | OLS<br>(9) | Poisson<br>(10) | OLS<br>(11) | Poisson<br>(12) |
| --- | --- | --- | --- | --- | --- | --- | --- | --- | --- | --- | --- | --- |
| 60+ adults (lag 6) | -0.53***<br>(0.13) | -0.67***<br>(0.12) |  |  |  |  |  |  |  |  |  |  |
| High-risk adults (lag 6) | -0.03<br>(0.14) | -0.12<br>(0.12) |  |  |  |  |  |  |  |  |  |  |
| Essential workers (lag 6) | -0.41***<br>(0.13) | -0.34<br>(0.26) |  |  |  |  |  |  |  |  |  |  |
| K-12 employees (lag 6) | 0.12<br>(0.13) | 0.16<br>(0.19) |  |  |  |  |  |  |  |  |  |  |
| 60+ adults (lag 7) |  |  | -0.59***<br>(0.12) | -0.68***<br>(0.12) |  |  |  |  |  |  |  |  |
| High-risk adults (lag 7) |  |  | -0.14<br>(0.13) | -0.23**<br>(0.13) |  |  |  |  |  |  |  |  |
| Essential workers (lag 7) |  |  | -0.42***<br>(0.13) | -0.31<br>(0.28) |  |  |  |  |  |  |  |  |
| K-12 employees (lag 7) |  |  | 0.02<br>(0.13) | 0.05<br>(0.22) |  |  |  |  |  |  |  |  |
| 60+ adults (lag 8) |  |  |  |  | -0.62***<br>(0.11) | -0.65***<br>(0.11) |  |  |  |  |  |  |
| High-risk adults (lag 8) |  |  |  |  | -0.27**<br>(0.13) | -0.31***<br>(0.14) |  |  |  |  |  |  |
| Essential workers (lag 8) |  |  |  |  | -0.40***<br>(0.13) | -0.26<br>(0.28) |  |  |  |  |  |  |
| K-12 employees (lag 8) |  |  |  |  | -0.08<br>(0.14) | -0.00<br>(0.24) |  |  |  |  |  |  |
| 60+ adults (lag 9) |  |  |  |  |  |  | -0.61***<br>(0.10) | -0.58***<br>(0.13) |  |  |  |  |
| High-risk adults (lag 9) |  |  |  |  |  |  | -0.37***<br>(0.13) | -0.36***<br>(0.15) |  |  |  |  |
| Essential workers (lag 9) |  |  |  |  |  |  | -0.33***<br>(0.13) | -0.16<br>(0.27) |  |  |  |  |
| K-12 employees (lag 9) |  |  |  |  |  |  | -0.22*<br>(0.14) | -0.09<br>(0.27) |  |  |  |  |
| 60+ adults (lag 10) |  |  |  |  |  |  |  |  | -0.55***<br>(0.10) | -0.45***<br>(0.15) |  |  |
| High-risk adults (lag 10) |  |  |  |  |  |  |  |  | -0.44***<br>(0.12) | -0.41***<br>(0.15) |  |  |
| Essential workers (lag 10) |  |  |  |  |  |  |  |  | -0.22*<br>(0.13) | -0.04<br>(0.26) |  |  |
| K-12 employees (lag 10) |  |  |  |  |  |  |  |  | -0.33***<br>(0.14) | -0.15<br>(0.29) |  |  |
| 60+ adults (lag 11) |  |  |  |  |  |  |  |  |  |  | -0.41***<br>(0.11) | -0.24*<br>(0.16) |
| High-risk adults (lag 11) |  |  |  |  |  |  |  |  |  |  | -0.46***<br>(0.11) | -0.41***<br>(0.14) |
| Essential workers (lag 11) |  |  |  |  |  |  |  |  |  |  | -0.06<br>(0.14) | 0.09<br>(0.24) |
| K-12 employees (lag 11) |  |  |  |  |  |  |  |  |  |  | -0.43***<br>(0.14) | -0.24<br>(0.29) |
| F-tests equality of coefficients: |  |  |  |  |  |  |  |  |  |  |  |  |
| 60+ = essential | 0.319 | 0.029 | 0.085 | 0.002 | 0.015 | 0.026 | 0.002 | 0.039 | 0.001 | 0.096 | 0.013 | 0.266 |
| 60+ = K-12 | 0.000 | 0.000 | 0.000 | 0.000 | 0.000 | 0.000 | 0.000 | 0.006 | 0.023 | 0.142 | 0.781 | 0.995 |
| 60+ = high-risk | 0.002 | 0.000 | 0.001 | 0.000 | 0.001 | 0.001 | 0.008 | 0.069 | 0.207 | 0.763 | 0.573 | 0.290 |
| essential = K-12 | 0.002 | 0.130 | 0.008 | 0.321 | 0.041 | 0.495 | 0.463 | 0.870 | 0.481 | 0.787 | 0.028 | 0.441 |
| essential = high-risk | 0.021 | 0.251 | 0.073 | 0.694 | 0.333 | 0.827 | 0.766 | 0.357 | 0.112 | 0.096 | 0.008 | 0.025 |
| K-12 = high-risk | 0.483 | 0.291 | 0.419 | 0.312 | 0.292 | 0.291 | 0.296 | 0.360 | 0.377 | 0.63 | 0.775 | 0.489 |
| Observations | 1,989 | 1,989 | 1,989 | 1,989 | 1,989 | 1,989 | 1,989 | 1,989 | 1,989 | 1,989 | 1,989 | 1,989 |
| (Pseudo-)R<sup>2</sup> | 0.625 | 0.740 | 0.641 | 0.741 | 0.654 | 0.740 | 0.658 | 0.736 | 0.648 | 0.732 | 0.634 | 0.729 |

Note: Robust standard errors clustered at the state level are displayed in parentheses below each respective coefficient; \*\*\* p<0.01, \*\* p<0.05, \* p<0.1 The interpretation of $100 \times \beta$ is the percentage change in cases in the state-week associated with a change of the regressor from 0 to 1.

**Table S5:** Prioritizations strategy regressed against share of population.

|  | Linear probability model |  |  |  | Logit (marginal effects) |  |  |  |
| --- | --- | --- | --- | --- | --- | --- | --- | --- |
|  | 60+<br>first<br>(1) | High-risk<br>first<br>(2) | Essential<br>first<br>(3) | K-12<br>first<br>(4) | 60+<br>first<br>(1) | High-risk<br>first<br>(2) | Essential<br>first<br>(3) | K-12<br>first<br>(4) |
| % 60+ adults (lag 6) | 0.00295<br>(0.0152) | -0.00221<br>(0.0276) | 0.00897<br>(0.0186) | -0.00363<br>(0.0271) | 0.00282<br>(0.0130) | -0.0102<br>(0.0289) | 0.00358<br>(0.0179) | -0.00870<br>(0.0282) |
| % High-risk adults (lag 6) | 0.0104<br>(0.00976) | 0.00709<br>(0.0169) | -0.00271<br>(0.0121) | 0.0116<br>(0.0170) | 0.0175**<br>(0.00789) | 0.00562<br>(0.0181) | -0.00559<br>(0.0119) | 0.00726<br>(0.0178) |
| % Essential workers (lag 6) | -0.00897<br>(0.00792) | -0.0105<br>(0.00926) | 0.00614<br>(0.00753) | 0.0132<br>(0.00985) | -0.0235**<br>(0.0119) | -0.0158<br>(0.0123) | 0.00258<br>(0.00642) | 0.00911<br>(0.0113) |
| % K-12 employees (lag 6) | -0.00597<br>(0.0167) | 0.0364<br>(0.0239) | -0.0105<br>(0.0207) | -0.0218<br>(0.0238) | -0.0100<br>(0.0188) | 0.0365<br>(0.0282) | -0.0122<br>(0.0188) | -0.0245<br>(0.0258) |
| F-tests equality of coefficients: |  |  |  |  |  |  |  |  |
| 60+ = essential | 0.516 | 0.761 | 0.874 | 0.532 | 0.203 | 0.843 | 0.953 | 0.515 |
| 60+ = K-12 | 0.690 | 0.354 | 0.545 | 0.657 | 0.605 | 0.346 | 0.577 | 0.729 |
| 60+ = high-risk | 0.750 | 0.830 | 0.691 | 0.721 | 0.383 | 0.722 | 0.749 | 0.716 |
| essential = K-12 | 0.890 | 0.107 | 0.498 | 0.229 | 0.607 | 0.122 | 0.499 | 0.281 |
| essential = high-risk | 0.201 | 0.458 | 0.620 | 0.950 | 0.0243 | 0.449 | 0.620 | 0.945 |
| K-12 = high-risk | 0.435 | 0.311 | 0.749 | 0.250 | 0.158 | 0.314 | 0.776 | 0.273 |
| Observations | 51 | 51 | 51 | 51 | 51 | 51 | 51 | 51 |
| Mean(dep. var.) | 0.098 | 0.294 | 0.216 | 0.725 | 0.098 | 0.294 | 0.216 | 0.725 |
| (Pseudo-)R2 | 0.162 | 0.333 | 0.217 | 0.722 | 0.2601 | 0.0710 | 0.0081 | 0.0665 |

Note: Robust standard errors clustered at the state level are displayed in parentheses below each respective coefficient; \*\*\* p<0.01, \*\* p<0.05, \* p<0.1

**Fig. S1:**
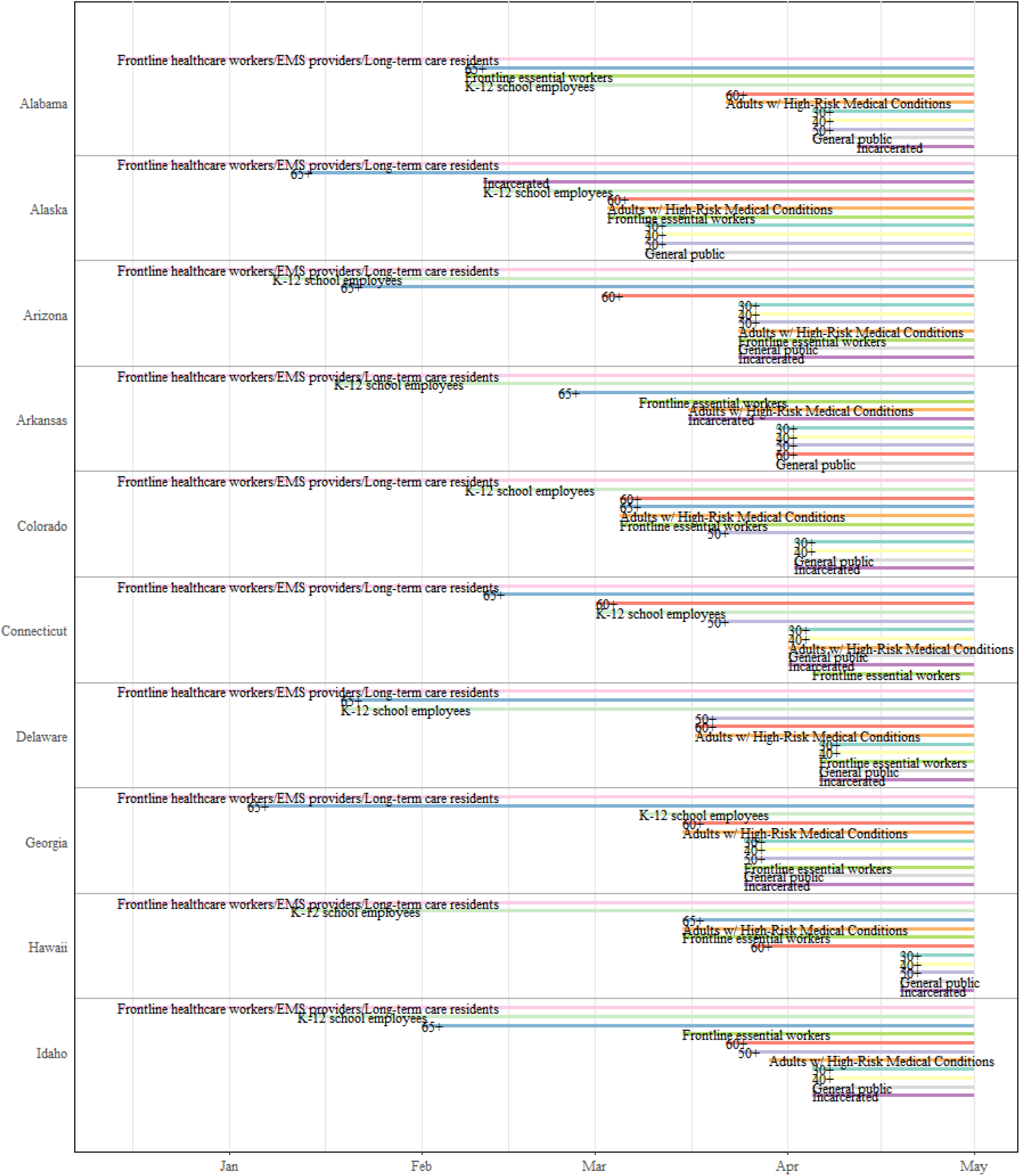
The timing of eligibility for Covid-19 vaccination in U.S. states. Each of these four states started their vaccination campaign on 14 December 2020 by vaccinating frontline healthcare workers, emergency medical service (EMS) providers, and long-term care residents. The ‘65+’ label corresponds to adults ages 65+ (with analogous labels for the other age groups). The eligibility start date was the same for the 65+, 70+, 75+ and 80+ groups in the four states shown. ‘Frontline essential workers’ are essential workers that are most likely at highest risk for work-related exposure to SARS-CoV-2, because their work-related duties must be performed on-site and involve being in close proximity ($<$6 feet) to the public or to coworkers. This group is proxied by grocery store workers, and includes firefighters, law enforcement and public safety personnel, correctional staff, U.S. Postal Service workers, food and agricultural workers, and manufacturing workers.

**Fig. S2:**
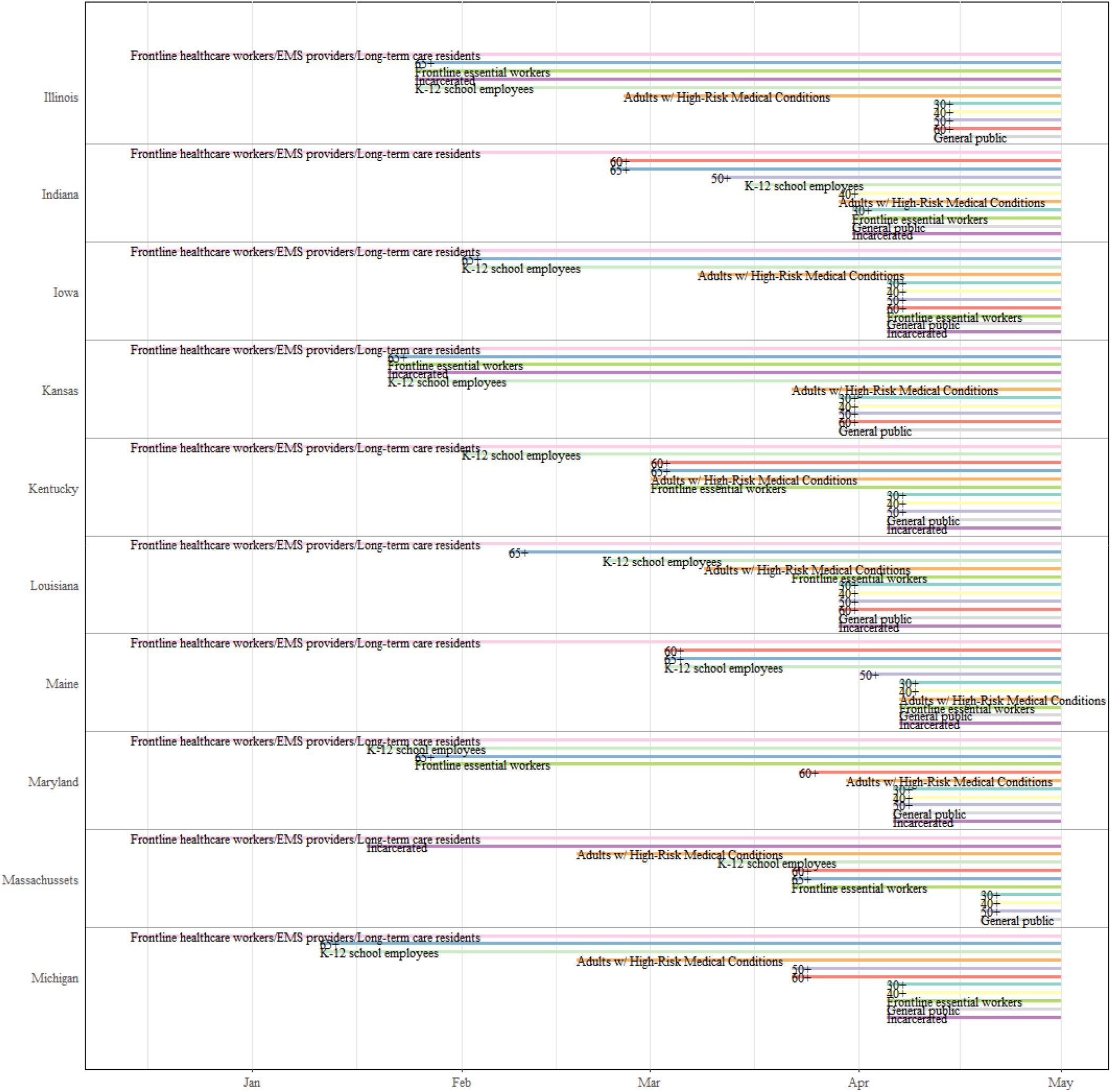
The timing of eligibility for Covid-19 vaccination in U.S. states. Each of these four states started their vaccination campaign on 14 December 2020 by vaccinating frontline healthcare workers, emergency medical service (EMS) providers, and long-term care residents. The ‘65+’ label corresponds to adults ages 65+ (with analogous labels for the other age groups). The eligibility start date was the same for the 65+, 70+, 75+ and 80+ groups in the four states shown. ‘Frontline essential workers’ are essential workers that are most likely at highest risk for work-related exposure to SARS-CoV-2, because their work-related duties must be performed on-site and involve being in close proximity ($<$6 feet) to the public or to coworkers. This group is proxied by grocery store workers, and includes firefighters, law enforcement and public safety personnel, correctional staff, U.S. Postal Service workers, food and agricultural workers, and manufacturing workers.

**Fig. S3:**
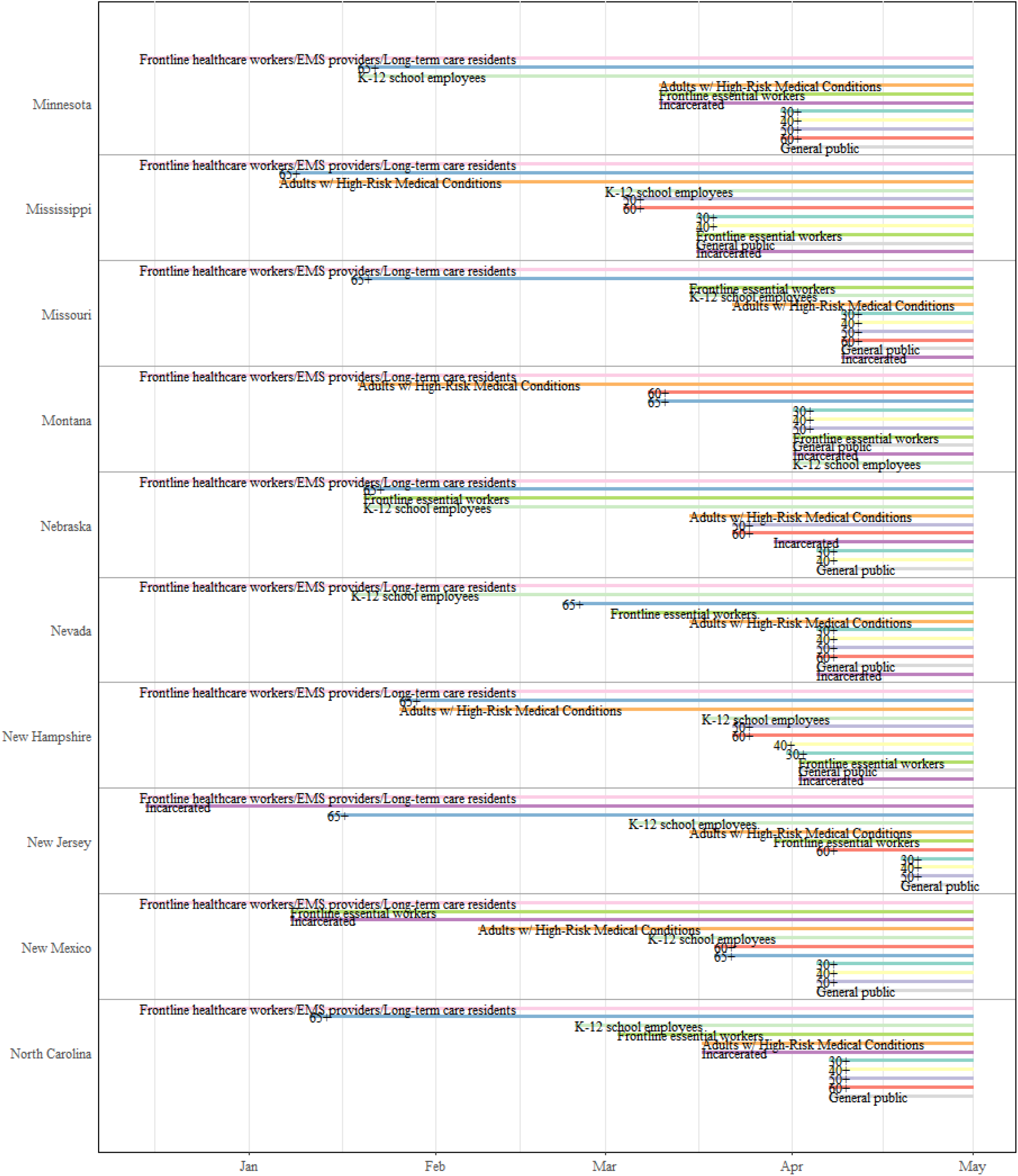
The timing of eligibility for Covid-19 vaccination in U.S. states. Each of these four states started their vaccination campaign on 14 December 2020 by vaccinating frontline healthcare workers, emergency medical service (EMS) providers, and long-term care residents. The ‘65+’ label corresponds to adults ages 65+ (with analogous labels for the other age groups). The eligibility start date was the same for the 65+, 70+, 75+ and 80+ groups in the four states shown. ‘Frontline essential workers’ are essential workers that are most likely at highest risk for work-related exposure to SARS-CoV-2, because their work-related duties must be performed on-site and involve being in close proximity ($<$6 feet) to the public or to coworkers. This group is proxied by grocery store workers, and includes firefighters, law enforcement and public safety personnel, correctional staff, U.S. Postal Service workers, food and agricultural workers, and manufacturing workers.

**Fig. S4:**
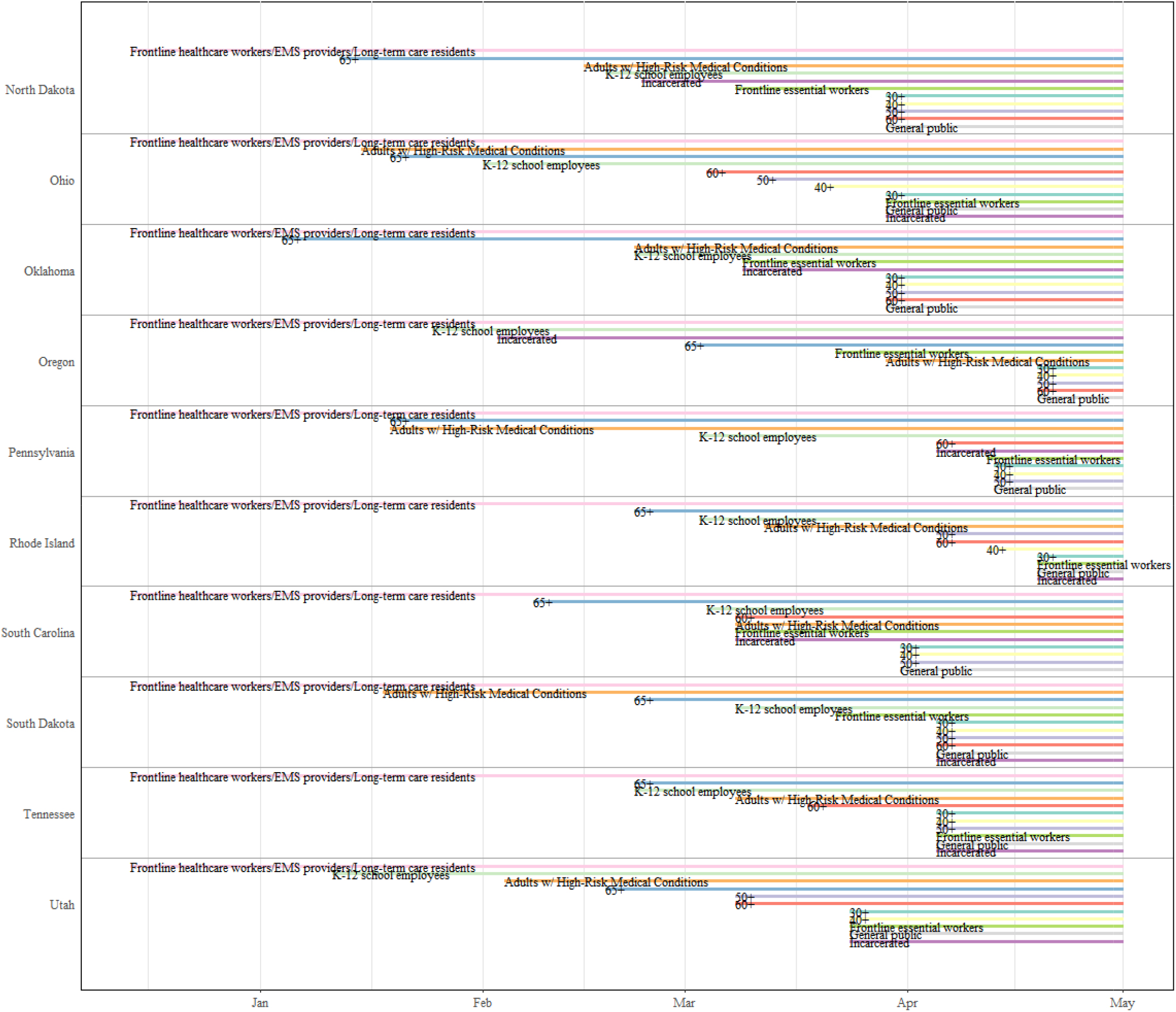
The timing of eligibility for Covid-19 vaccination in U.S. states. Each of these four states started their vaccination campaign on 14 December 2020 by vaccinating frontline healthcare workers, emergency medical service (EMS) providers, and long-term care residents. The ‘65+’ label corresponds to adults ages 65+ (with analogous labels for the other age groups). The eligibility start date was the same for the 65+, 70+, 75+ and 80+ groups in the four states shown. ‘Frontline essential workers’ are essential workers that are most likely at highest risk for work-related exposure to SARS-CoV-2, because their work-related duties must be performed on-site and involve being in close proximity ($<$6 feet) to the public or to coworkers. This group is proxied by grocery store workers, and includes firefighters, law enforcement and public safety personnel, correctional staff, U.S. Postal Service workers, food and agricultural workers, and manufacturing workers.

**Fig. S5:**
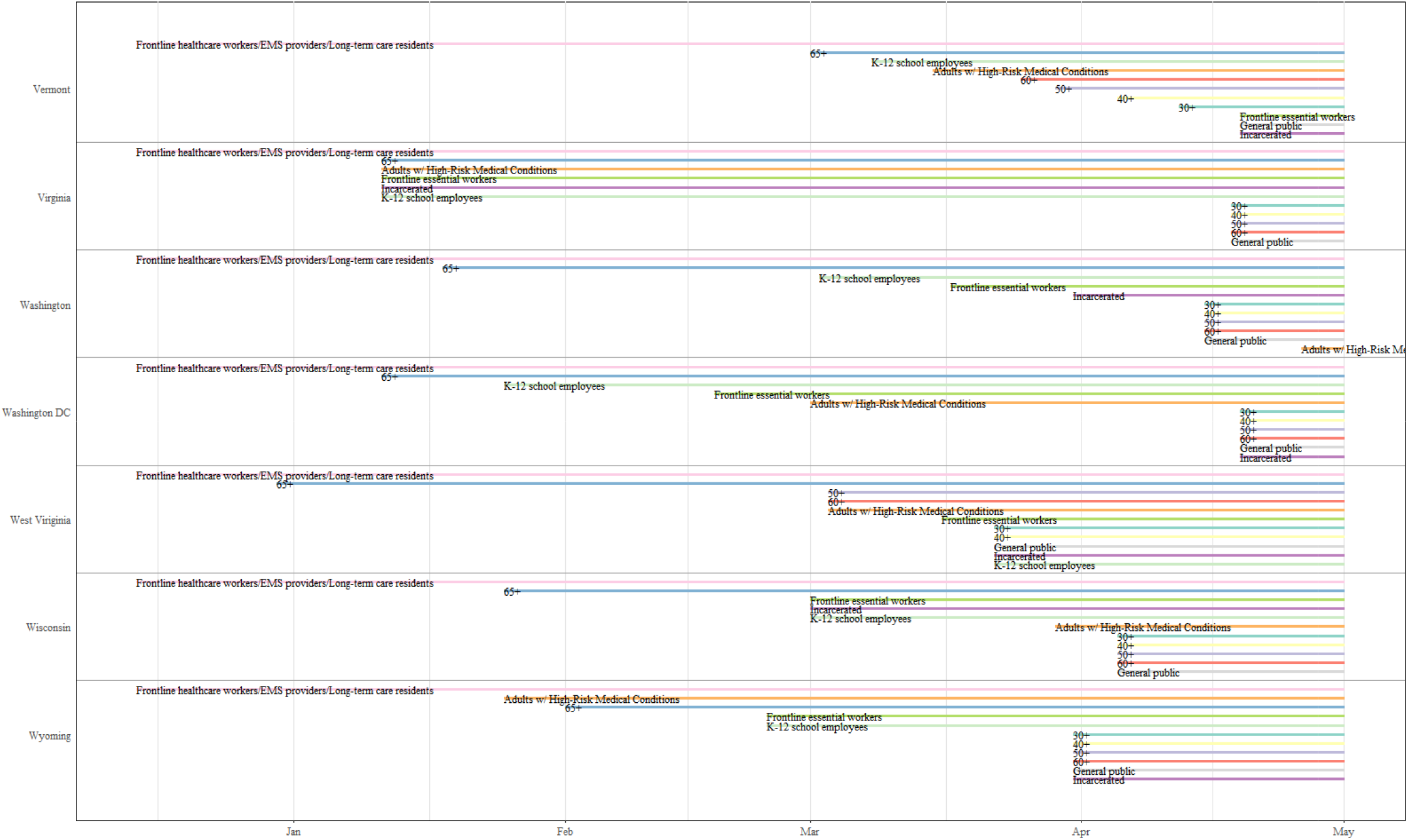
The timing of eligibility for Covid-19 vaccination in U.S. states. Each of these four states started their vaccination campaign on 14 December 2020 by vaccinating frontline healthcare workers, emergency medical service (EMS) providers, and long-term care residents. The ‘65+’ label corresponds to adults ages 65+ (with analogous labels for the other age groups). The eligibility start date was the same for the 65+, 70+, 75+ and 80+ groups in the four states shown. ‘Frontline essential workers’ are essential workers that are most likely at highest risk for work-related exposure to SARS-CoV-2, because their work-related duties must be performed on-site and involve being in close proximity ($<$6 feet) to the public or to coworkers. This group is proxied by grocery store workers, and includes firefighters, law enforcement and public safety personnel, correctional staff, U.S. Postal Service workers, food and agricultural workers, and manufacturing workers.

**Fig. S6:**
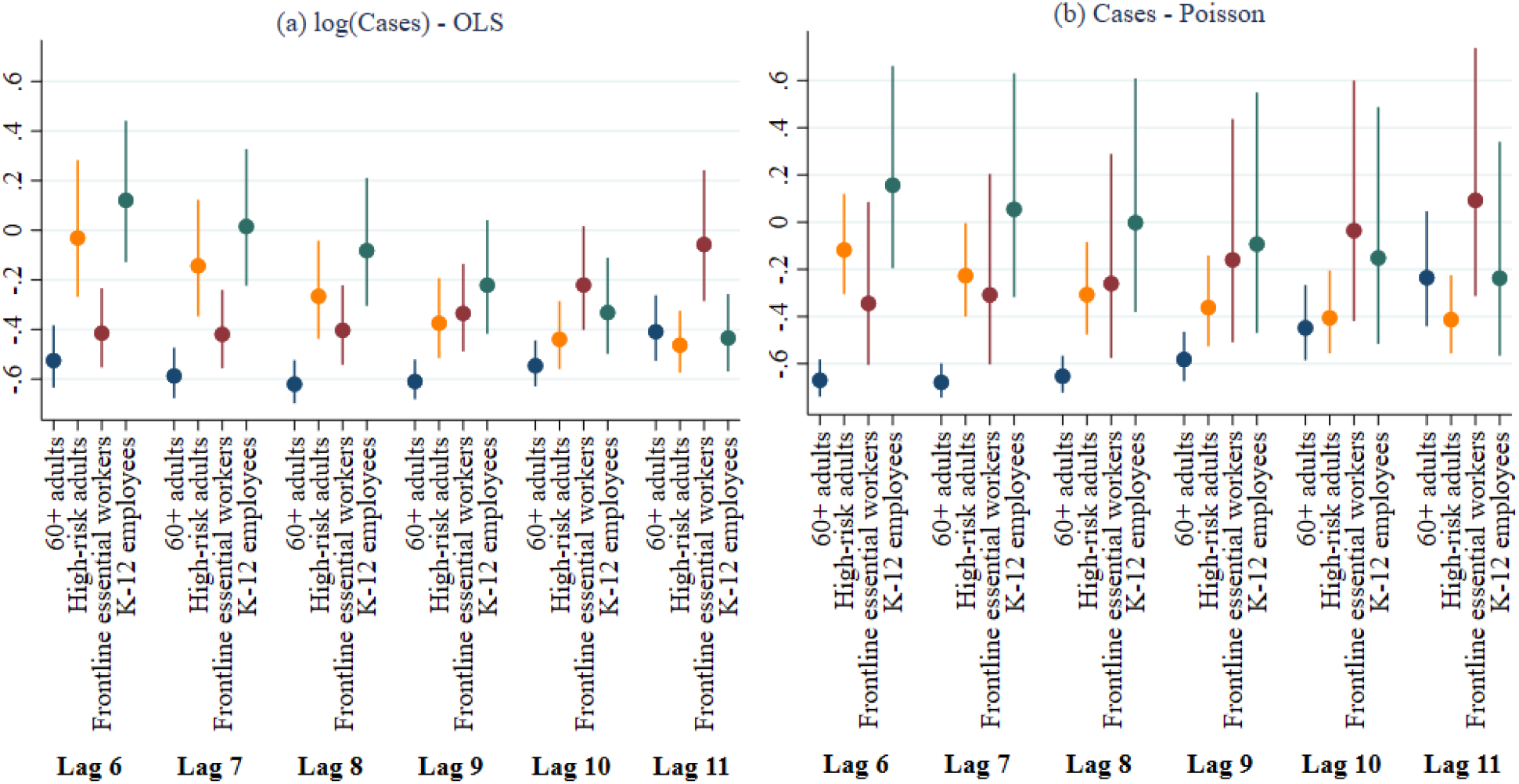
In regressions of Covid-19 cases, the coefficients are plotted of the indicators for whether a ACIP group was eligible in a state, with a lag of 6-11 weeks. Panel (a) displays estimates of the coefficients and 95% confidence intervals of log-linear models estimated by OLS (after the transformation (exp(β) - 1)), and panel (b) displays coefficient estimates and 95% confidence intervals of Poisson regressions. In both panels, 100 times the displayed (transformed) coefficients can be interpreted as the percentage change relative to the benchmark where neither of the four groups is eligible for vaccination in that week/lag.

